# Distance to white matter trajectories is associated with treatment response to internal capsule deep brain stimulation in treatment-resistant depression

**DOI:** 10.1101/19008268

**Authors:** Luka C. Liebrand, Samuel J. Natarajan, Matthan W.A. Caan, P. Rick Schuurman, Pepijn van den Munckhof, Bart de Kwaasteniet, Judy Luigjes, Isidoor O. Bergfeld, Damiaan Denys, Guido A. van Wingen

## Abstract

**Objective:** Deep brain stimulation (DBS) is an innovative treatment for treatment-resistant depression. DBS is usually targeted at specific anatomical landmarks, with patients responding to DBS in approximately 50% of cases. Attention has recently shifted to white matter tracts to explain DBS response, with initial open-label trials targeting white matter tracts yielding much higher response rates (>70%).

**Methods:** We associated distance to individual white matter tracts around the stimulation target in the ventral anterior limb of the internal capsule to treatment response. We performed diffusion magnetic resonance tractography of the superolateral branch of the medial forebrain bundle and the anterior thalamic radiation in fourteen patients that participated in our randomized clinical trial. We combined the tract reconstructions with the postoperative images to identify the DBS leads and estimated the distance between tracts and leads, which we subsequently associated with treatment response.

**Results:** Stimulation closer to both tracts was significantly correlated to a larger symptom decrease (r=0.61, p=0.02), suggesting that stimulation more proximal to the tracts was beneficial. There was no difference in lead placement with respect to anatomical landmarks, which could mean that differences in treatment response were driven by individual differences in white matter anatomy.

**Conclusions:** Our results suggest that deep brain stimulation of the ventral anterior limb of the internal capsule could benefit from targeting white matter bundles. We recommend acquiring diffusion magnetic resonance data for each individual patient.

## INTRODUCTION

Deep brain stimulation (DBS) is an innovative last-resort treatment for treatment-resistant depression (TRD). Patients in DBS trials usually failed to respond to multiple adequate treatments, including antidepressants and electroconvulsive therapy. Approximately 10-15% percent of patients with depression has a severe level of treatment-resistant depression.[1] DBS studies have shown promising results with half of patients responding to DBS. However, results of randomized controlled trials have been mixed, with some showing large differences between active and sham DBS,[2–4] and others failing to find differences.[5,6]

Different brain regions have been targeted for TRD, including the subcallosal cingulate,[7] anterior limb of the internal capsule,[2] the ventral capsule/ventral striatum,[8] and nucleus accumbens.[9] The mechanism of action of DBS seems to be that it normalizes pathological network connectivity, [10] which has motivated specifically targeting white matter tracts that make up these networks.[11,12]

The most popular method for in-vivo reconstruction of white matter tracts is tractography in diffusion magnetic resonance imaging (dMRI) data. Several groups have reported retrospective or prospective open-label studies where they used tractography to refine surgical targets. In retrospective studies, the goal was often to determine whether proximity to, or activation of, white matter tracts is related to treatment response, whereas prospective studies aimed to exploit this knowledge by selectively targeting or avoiding one or more tracts.[13] In this retrospective study, we are interested in a relationship between tracts coursing through the ventral anterior limb of the internal capsule (ALIC) and treatment response.

The white matter anatomy of the ALIC has been shown to be well-ordered, but variable along individuals.[11,14–16] It was hypothesized that stimulation of disrupted dopaminergic connections from the ventral tegmental area (VTA) to the nucleus accumbens/striatum might be related to treatment response for TRD.[17] Research based on this hypothesis disentangled two important fiber pathways coursing through the ALIC: the anterior thalamic radiation (ATR), and the superolateral medial forebrain bundle (slMFB)[11]. The slMFB makes up the rostral part of the cortico-pontine connection between the VTA and prefrontal cortex, whereas the ATR originates in the anterior and dorsomedial thalamus, also connecting to the prefrontal cortex through the ALIC.

Stimulation of the slMFB, as the dopaminergic connection between the VTA and striatum, proposedly elicits response through normalization of striatal dopamine levels. This idea is in line with the finding that ALIC stimulation induced striatal dopamine release in patients with obsessive-compulsive disorder (OCD).[18] Taken together, this theoretical framework has resulted in the investigation of the slMFB as a stimulation target for TRD,[3,19,20] although closer to the VTA instead of in the ALIC.

Based on the work on sIMFB stimulation and our previous finding that proximity of stimulation to sIMFB was related to treatment response in OCD,[21] we hypothesize that stimulation more proximal to the slMFB within the ALIC is also beneficial for treatment response in TRD. However, a possible role of the ATR and thalamus cannot be ruled out, given reported structural changes within the thalamus,[22] and hyperactivity of the pulvinar nucleus in MDD patients.[23] Therefore, here we use tractography to establish whether there is a relationship between proximity of stimulation to the slMFB and ATR with respect to treatment outcome. The findings could have a direct clinical impact by refining the surgical target in future cases and could lead to reevaluation of DBS lead placement in our current non-responders.

## METHODS

### Participants

The data for this study were acquired as part of the clinical trial published by Bergfeld and colleagues (2016).[2] This trial was a collaboration between the Academic Medical Center (AMC) in Amsterdam, and the St. Elizabeth Hospital in Tilburg, both in the Netherlands, and was approved by the medical ethics committees of both hospitals.

Patients (aged 18 to 65 years) had a primary diagnosis of major depressive disorder (MDD), with an illness duration of >2 years, a score of ≥18 on the 17-item Hamilton depression rating scale (HAM-D), and a global assessment of function Score of ≤45. Treatment resistance was defined as a failure of response to: two classes of second-generation antidepressants; two single trials of a tricyclic antidepressant (with and without lithium augmentation, respectively); one trial of a monoamine oxidase inhibitor; and bilateral electroconvulsive therapy for ≥6 sessions. Additionally, patients had to have an IQ of >80 and be eligible for surgery. Exclusion criteria were schizophrenia, psychosis unrelated to MDD, bipolar disorder, recent substance abuse (i.e. within the past 6 months), antisocial personality disorder, Parkinson’s disease, dementia, epilepsy, tic disorder, and pregnancy. In addition to abovementioned criteria, sufficient quality imaging data – particularly dMRI scans suitable for tractography – were necessary for inclusion into present study.

### DBS surgery and treatment

Structural and diffusion-weighted MRI scans were made at 3T at baseline. Imaging details are described in the “*Imaging*” section. A stereotactic frame was attached to the patient on the morning of surgery. The patient was subsequently scanned at 1.5T to express the surgical planning in stereotactic coordinates. The neurosurgeon performed the surgical planning in SurgiPlan (Elekta AB, Stockholm, Sweden) according to standard stereotactic procedures.[24] In short, the following coordinates relative to the intercommissural line were the starting point of surgical planning: 3 mm anterior to the anterior border of the anterior commissure, 7 mm lateral to the midline, and 4 mm inferior to the intercommissural line. From there, the bilateral targets were refined with respect to the nucleus accumbens (NAc) and ALIC, so that the deepest of four contacts was placed in the NAc and the remaining contacts were placed in the ventral ALIC. Electrodes (model 3389, Medtronic, Minneapolis, MN, USA) with 1.5 mm contacts and 0.5 mm interspace were placed along a sagittal angle of approximately 75° to the intercommissural line, and a coronal angle following the ALIC into the NAc. Directly after surgery, a computed tomography (CT) or 1.5T structural MRI scan was made to ensure correct lead placements.

Two weeks after implantation, the DBS device was switched on and the DBS settings optimization phase started. We refer to Bergfeld et al. (2016) for details.[2] Optimization was done open-label and lasted up to 52 weeks. For the present study, we were primarily interested in the relationship between the active contacts relative to tracts within the ALIC, and treatment outcome. We used the optimized DBS settings from before the randomized cross-over period. Choosing the HAM-D scores after optimization ensured that patients had stable (and optimized) stimulation parameters, and that the cross-over period, which was associated with significant worsening of symptoms, did not affect our results. Treatment response was measured by the percentage difference in HAM-D scores between baseline and post-optimization follow-up.

### Imaging: acquisition

Our aim was to represent individual patients’ white matter tracts relative to the electrodes. For this reason, we combined the post-operative CT scans with tractography results from the pre-operative 3T dMRI scans in each patient’s native structural space (i.e. pre-operative 1.5T T1-weighted scans). This approach has the benefit compared to an atlas-based approach that it retained as much individual information as possible, thereby allowing to better assess individual differences. A schematic overview of this procedure is given in figure 1.

**Figure 1.**
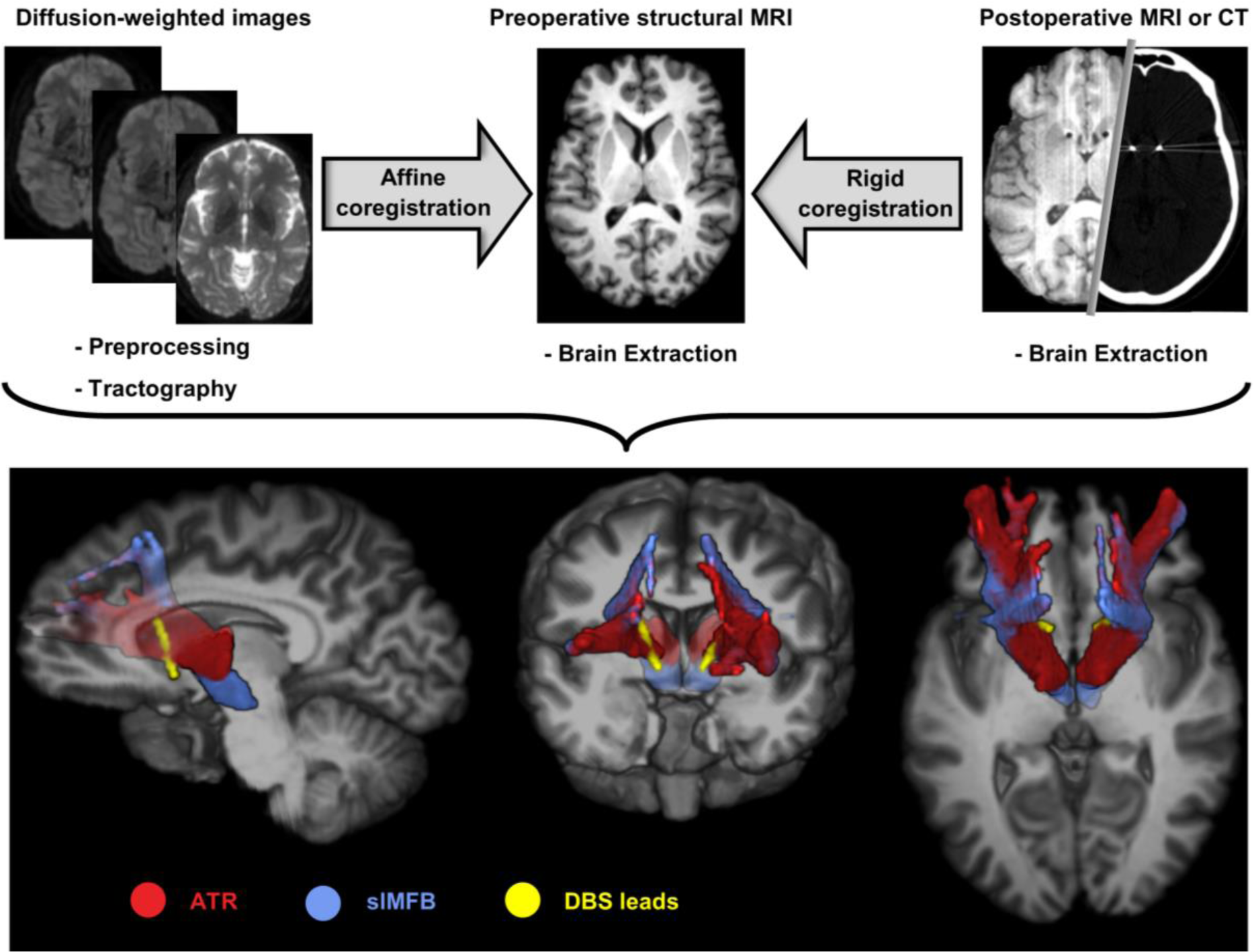
Schematic overview of analysis pipeline. **(Top row)** The preprocessed diffusion data were used to generate tractography results. The tractography results were affinely coregistered to the brain-extracted preoperative structural scan. The postoperative scan was rigidly coregistered to add the lead localization. **(Bottom row)** 3D-rendering of one patient’s structural scan, overlaid with the reconstructed anterior thalamic radiation (ATR), superolateral medial forebrain bundle (slMFB), and DBS leads. Views are as follows: **(left)** sagittal (right hemisphere), **(middle)** coronal view (front-facing), and **(right)** axial view (top-down).

All 3T scans were made at a Philips Ingenia scanner (Philips Medical Systems, Best, The Netherlands) equipped with a 16-channel phased-array headcoil. The T1-weighted scans were sagittally acquired on a 1.5T Siemens Avanto scanner, with a 3D inversion-recovery sequence with 0.9 × 0.9 × 1.2 mm^3^ voxel size and 256×256 ×182 matrix size. The diffusion-weighted scans were acquired according to a 2D Stejskal-Tanner spin-echo sequence, with 2.0^3^ mm^3^ resolution, 112×112×70 matrix, 32 non-collinear directions with b = 600 s/mm^2^ and one b = 0 s/mm^2^, TE = 60 ms, TR = 6770 ms. Post-operative MRI scans were made at a 1.5T, at a resolution of 1.0 × 1.0 × 1.0 mm^3^, 192×256×256 matrix size, TE = 3 ms, TR = 1900 ms. The CT scans had a resolution of 0.45 × 0.45 × 1.0 mm^3^ and 512×512×162 matrix size.

### Imaging: (pre)processing

The preprocessing for the structural MRI scans consisted of brain extraction with FSL’s *bet* toolbox (FMRIB Software Library, version 5.0.10, https://fsl.fmrib.ox.ac.uk/). The post-operative CT scans were brain extracted with a custom Matlab script (version R2016a, The Mathworks, Natick, MA). The postoperative T1 and CT scans were rigidly coregistered to the preoperative T1 scans with FSL’s Flirt tool.

Preprocessing of dMRI data consisted of eddy current and movement correction with FSL’s eddy correct tool, which coregistered all diffusion-weighted images to the b0 image. The b-vectors were rotated accordingly.[25] An in-house developed Matlab script was used to correct for ringing artefacts. We calculated the affine transformations between structural and diffusion space (i.e. preoperative T1 and b0 image, respectively) with ANTS symmetric diffeomorphic registration (Advanced Normalization Toolbox, version 2.1.0, http://stnava.github.io/ANTs/;).[26] Voxelwise diffusion orientations were estimated with a model that accounts for crossing fibers (FSL’s BedpostX;).[27]

### Tractography

In this study, we were interested in reconstructing the slMFB and ATR. Tractography seeds were drawn bilaterally in the VTA for the slMFB, and anterior thalamus for the ATR, with a common waypoint in the ALIC, according to the work by Coenen et al. (2012).[11] Probabilistic tractography was performed with FSL’s probtrackx (default parameters). Tracking results were visually inspected and tractography seeds were refined if necessary. Finally, tractography results were transformed to structural space according to the earlier calculated transformations.

### Distance from tracts to contacts

Before calculating the distances between the contacts and the tracts, the location of the active contacts was determined. This was done by tracing a path along the center of the electrode artefact on the CT scan, starting from the tip located in the NAc. All contacts were labeled and visually checked for accuracy. In subsequent stages, only the active contacts for each patient were used. The shortest distances between the electrode contacts and tracts were calculated in 3D in Matlab, with a heuristically determined threshold of 34% that yielded the optimal distribution of distances for statistical analysis. We estimated the average distance *d* from contact to tract for both tracts, 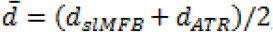, and the difference between distances from contact to tract of both tracts, Δ*d* = *d*_*slMFB*_−*d*_*ATR*_. Here, *d*_*slMFB*_ and *d*_*ATR*_ represent the (average of left and right) distance to respectively the slMFB and the ATR. If multiple contacts were active, the distance to the closest contact was chosen.

### Statistical analysis

We correlated the percentage change in HAM-D scores between baseline and follow-up for each patient with the average distance of the contacts to both bundles 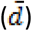 (i.e. main effect term), and the differential distance to the bundles (Δ*d*) (i.e. interaction term). Because of the apparent non-Gaussian distribution of the data, we calculated the non-parametric Spearman’s ranked correlation.

## RESULTS

Out of a cohort of 25 patients, ten patients did not have a complete dataset consisting of preoperative T1 and dMRI scans, and a postoperative T1 or CT scan. One patient’s dMRI scan suffered from large movement artefacts and was excluded. This resulted in a total of 14 subjects of whom we had a complete dataset of sufficient quality for inclusion into this study. The treatment response in this cohort was on average 7.4 points (−33%) on the 17-point HAM-D scale, with seven patients being responders (classified as at least 50% decrease in symptom scores). An overview of treatment response and stimulation settings is shown in Table 1.

**Table 1.** Patient response and DBS settings. Overview of treatment response and stimulation settings of all included patients.

| Patient | Change in HAM-D (%) | Responder (Yes/No) | Active contacts (Left)<br>Lowest-highest: 0-3 | Voltage (Left) (V) | Active contacts (Right)<br>Lowest-highest: 8-11 | Voltage (Right) (V) | Stim. frequency (Hz) | Pulse duration (μs) |
| --- | --- | --- | --- | --- | --- | --- | --- | --- |
| 01 | -87.5 | Y | 2, 3 | 5.0 | 10, 11 | 3.5 | 180 | 120 |
| 02 | +36.8 | N | 0, 1, 2 | 3.8 | 9, 10, 11 | 3.8 | 190 | 90 |
| 03 | -62.5 | Y | 2 | 4.3 | 10 | 4.3 | 180 | 90 |
| 04 | -77.8 | Y | 1, 2 | 5.5 | 9, 10 | 5.5 | 180 | 90 |
| 05 | +54.5 | N | 0, 1 | 5.5 | 9, 10 | 5.5 | 130 | 90 |
| 06 | -50.0 | Y | 2, 3 | 4.3 | 10, 11 | 4.3 | 180 | 90 |
| 07 | +6.3 | N | 1, 2 | 5.4 | 9, 10 | 5.4 | 180 | 90 |
| 08 | -72.7 | Y | 1, 2, 3 | 7.3 | 9, 10, 11 | 7.3 | 180 | 90 |
| 09 | -53.3 | Y | 1, 2 | 3.5 | 9, 10, 11 | 6.0 | 180 | 90 |
| 10 | -8.3 | N | 1, 2 | 5.0 | 9, 10 | 5.0 | 130 | 90 |
| 11 | +8.3 | N | 2, 3 | 6.7 | 10, 11 | 6.7 | 180 | 90 |
| 12 | -27.3 | N | 2, 3 | 2.5 | 10, 11 | 2.5 | 130 | 60 |
| 13 | -83.3 | Y | 1, 2 | 5.4 | 9, 10 | 5.4 | 180 | 90 |
| 14 | -30.4 | N | 1 | 5.2 | 9 | 5.2 | 130 | 60 |

For all included subjects, we were able to reconstruct both tracts of interest (slMFB and ATR). As expected, the reconstructions of the slMFB and ATR could be clearly distinguished from their respective starting points in the VTA and anterior thalamus, up to the ALIC, where they were often laterally-medially organized, and slightly overlapping. Finally, both tracts terminated in the prefrontal cortex. The tracts followed roughly the same trajectory and respective organization in each individual, although there were differences in the exact trajectory. In order to give an impression of the variability within the ALIC, an overview of tractography results is shown in figure 2. For most subjects, the trajectory of the slMFB and ATR were located dorsally in the ALIC, and, consequently, also with respect to the DBS contacts. This is reflected in the relatively high average distances from the active contacts to both bundles 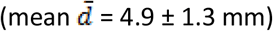, as can be seen in figure 3.

**Figure 2.**
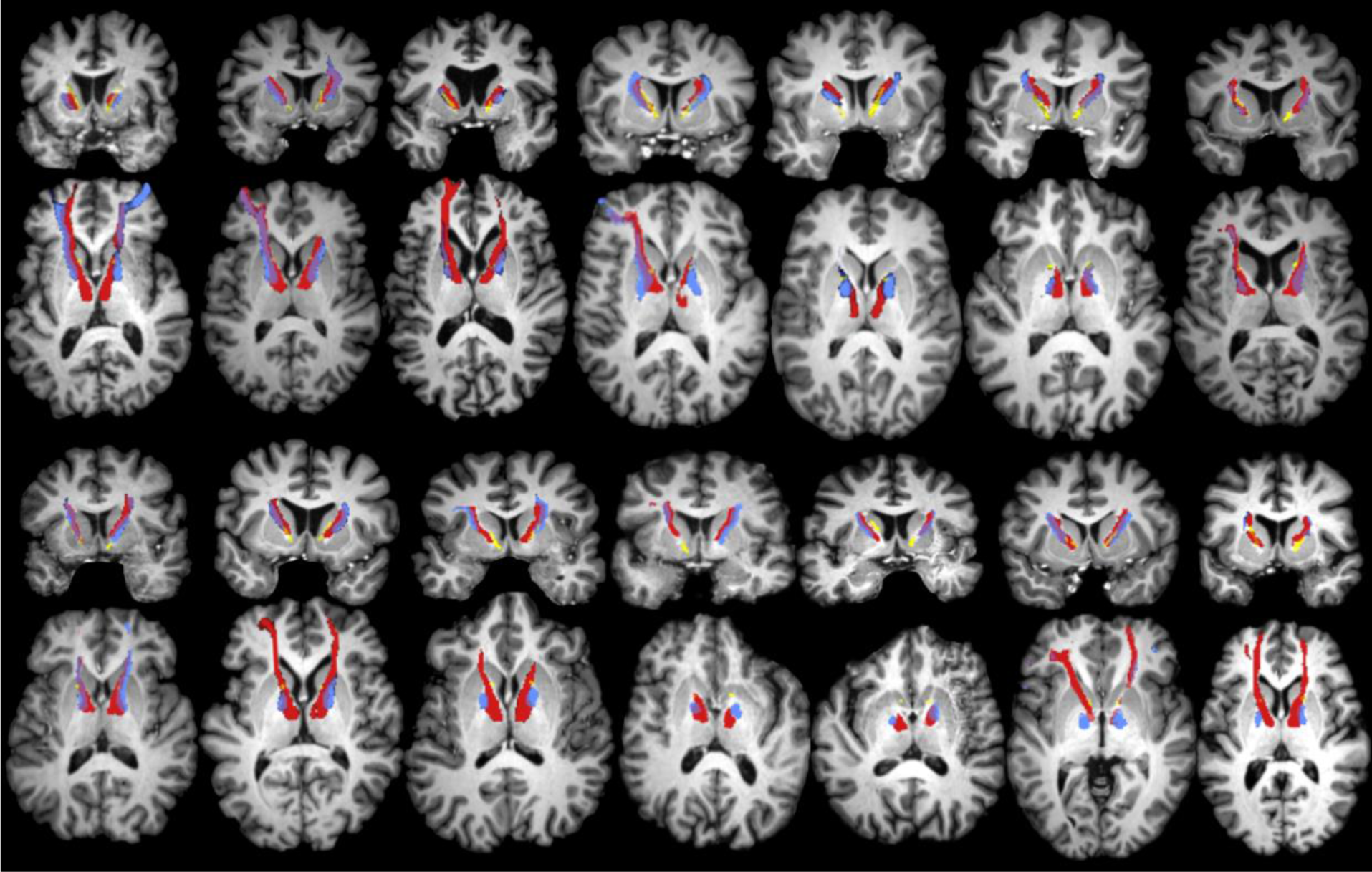
Overview of tractography results for all patients. Coronal and axial views of reconstructed anterior thalamic radiation (ATR), superolateral medial forebrain bundle (slMFB), and DBS leads, for all 14 subjects included in this study. Each coronal view corresponds to the axial view directly below. Colour coding is identical to Figure 1. It can be seen that the ATR is consistently medial to the slMFB within the anterior limb of the internal capsule (ALIC). For some subjects, the slMFB appears more dorsal in the ALIC than the ATR.

**Figure 3.**
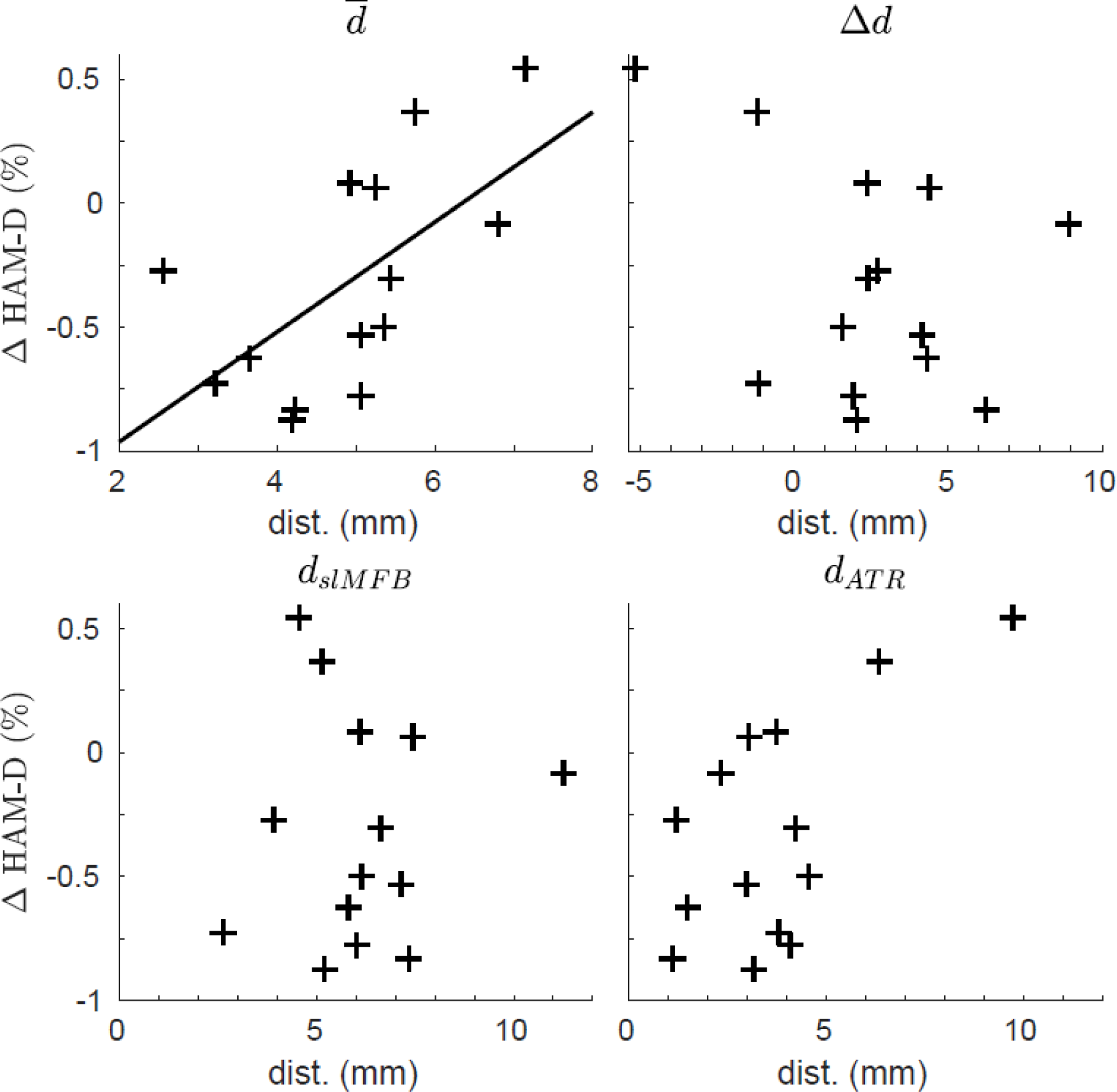
Distance from tracts to contacts associated with response. Scatter plots showing the relationship between distance of the anterior thalamic radiation (ATR) or superolateral medial forebrain bundle (slMFB) to the active contact, and treatment response (percentage change in symptoms). The different panels include **(top left)** the average distance to both bundles (main effect), **(top right)** the difference between distances (interaction term), **(bottom left)** relationship to slMFB only, and **(bottom right)** relationship to the ATR. Only the relationship between the average distance to both bundles and treatment response **(top left)** was significant (r = 0.61, p = 0.02), which is indicated by the line.

There was a significant relationship between average distance 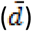 and percentage response (r = 0.61, p = 0.02). In contrast, there was no significant relationship between the differential distance (Δ*d*) and response (r = −0.20, p = 0.50). Post-hoc, we also related the distances from the active contacts to either the slMFB (r = −0.02, p = 0.96) or ATR (r = 0.39, p = 0.17), to the change in HAM-D, but these correlations were not significant.

### Stimulation site comparison

In addition to tractography in individual patient space, we show the overlap of individual stimulation sites after nonlinear transformation to MNI-space (figure 4). The stimulation sites show a high degree of overlap, suggesting that there was no difference in placement with respect to anatomical landmarks between responders and nonresponders.

**Figure 4.**
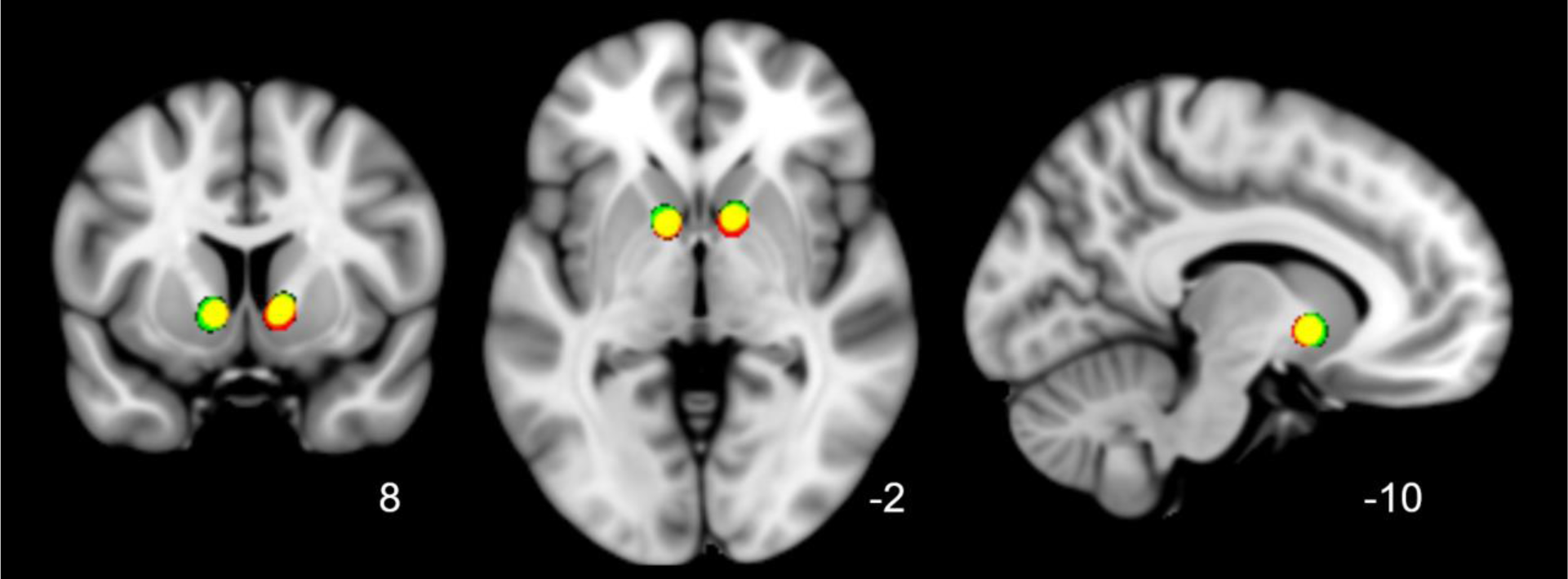
Overlap of active stimulation sites of (non)responders in standard (MNI) space. Transformed and smoothed (4 mm FWHM) stimulation sites of all subjects shown in standard MNI space (1 mm) with respective coronal, axial and sagittal views. Colour coding: responders (green), nonresponders (red), overlap (yellow). Stimulation sites of responders and nonresponders were all located in the ventral anterior limb of the internal capsule, directly above the nucleus accumbens, and almost completely overlapped. This suggests that differences in treatment outcome were unrelated to stimulation with respect to anatomical landmarks.

## DISCUSSION

In this work, we set out to determine whether the treatment outcome of DBS of the ventral ALIC for TRD was related to the stimulation’s proximity to the slMFB and ATR white matter tracts, using tractography to reconstruct their likely trajectories. On average, the tracts were located quite dorsally with respect to the stimulation site in the ventral ALIC, directly above the nucleus accumbens. By relating the distances of the slMFB and ATR to the active contacts to treatment response, we discovered that stimulation closer to both bundles was associated with better treatment outcome.

This result supports recent studies indicating the potential of tract stimulation in DBS for TRD, [3] and agrees with our finding in obsessive-compulsive disorder suggesting treatment response is related to tract proximity.[21] In addition, the large degree of overlap in stimulation sites suggests that treatment outcome does not depend on lead placement with respect to anatomical landmarks. The outcome of this study may be of clinical relevance, and prospective studies have to determine whether tractography-assisted surgical targeting in vALIC DBS for TRD is indeed beneficial. In addition, this result suggests that patients with limited clinical response might benefit from repositioning the leads.

Based on earlier work by others and our findings in OCD, we hypothesized that the slMFB would be the preferable target over the ATR in the ALIC. A prominent role for the slMFB is supported by recent promising results of slMFB stimulation close to the VTA, distant from the ATR.[3] Contrary to our expectations, there was no significant relationship to the individual proximity of either bundle and treatment outcome. The large distance between the leads with respect to both tracts might have made it difficult to differentiate each tract’s contribution to the treatment outcome. Therefore, given present findings, we cannot invalidate the hypothesis that the slMFB is the preferable target. However, we cannot rule out a potential role of the ATR in ALIC DBS for TRD either.

Little evidence points to the ATR as the optimal target in ALIC DBS for TRD, although a recent study did find a positive association between stimulation of frontothalamic connections in the ALIC and treatment outcome for OCD.[28] While structural changes in the thalamus,[22] and hyperactivity in the pulvinar nucleus have been reported in patients with MDD,[23] these are outside the context of DBS for TRD. Nevertheless, disruption of frontothalamic connectivity through stimulation of the ATR might have been (at least partly) responsible for improvement of depressive symptoms.[28]

Possible working mechanisms of slMFB stimulation have been proposed, suggesting an important role for the dopaminergic connections from the VTA to the striatum and prefrontal cortex.[11] This is supported by work showing ventral ALIC stimulation in OCD patients was associated with an increase in striatal dopamine.[18] The supposedly central role of the VTA has motivated stimulation of the slMFB much closer to the VTA, and away from the ALIC.[19] Interestingly, there is a possibility that the tract itself is the optimal target, relatively independent of where it is being stimulated, which could suggest a common network across different stimulation targets that underlies DBS response in TRD. Future research should focus on determining common prefrontal connections of the slMFB, ATR, and other DBS targets to confirm the existence of such a network. Moreover, since the ALIC is a white matter hub with many different prefrontal connections,[16,29] it may be useful to identify adjacent fiber connections and their relationship to treatment response and side effects in future studies.

### Limitations

This work is primarily limited by the number of subjects (N=14). Sample size is a limitation in most DBS studies for psychiatric indications, and our sample size is comparable to other tractography studies in this field. Nevertheless, care must be taken in interpretation of the results, and future studies should aim to replicate these findings, possibly pooling data of multiple centers using the same target to overcome the limited sample sizes inherent to psychiatric DBS. Even so, we were able to find an association between overall proximity of the slMFB and ATR to the active DBS contacts and treatment outcome. Our relatively straight-forward study design further facilitated interpretation of the results. We therefore believe that our sample size was sufficient for this study.

Another limitation lies in the retrospective character of this study. Surgical targeting during this study was based on anatomical landmarks, notably the nucleus accumbens, causing the stimulation site to be quite ventral within the ALIC. Although the resulting variability in distance between the contacts and tracts actually enabled the current study, the large distance made it more difficult to associate treatment response to stimulation of one tract specifically. In prospective studies, there can be much more control over the positioning of the electrodes with respect to the tracts, allowing a direct comparison between slMFB and ATR stimulation.

Finally, we were limited by the qualitative nature of tractography,[30] which makes it difficult to determine the volume of a tract. As a result, distances from tracts to the active contacts may not be exact, although we believe this does not undermine the validity of our results. Our findings do not dependent on precise distance but the variability in distance between subjects. We specifically avoided quantification of connectivity strength to and from our stimulation target, since tractography is unsuited for this.[31] By taking care in assessing our results, and using an easily interpretable method that can also be used for surgical planning, we believe that we have found a middle ground between usability and prudence.

### Outlook

Based on our results, we recommend and will incorporate tractography-guided surgical planning in order to target the slMFB and ATR within the internal capsule for TRD. It is probable that within the ALIC, the slMFB is the optimal target, although future studies stimulating closer to both targets should be done to be able to discern the slMFB and ATR. Even for other DBS targets and indications, we recommend collecting dMRI data, in order to perform retrospective studies to elucidate the potential role of white matter tracts in response.

## CONCLUSION

In this work, we show that stimulation closer to the slMFB and ATR in the ventral ALIC is associated with better treatment outcome in TRD. We were not able to distinguish between individual contributions of slMFB and ATR stimulation, probably due to stimulation in this study being delivered too ventrally with respect to these bundles. There seems to be no relationship between lead placement with respect to anatomical landmarks and treatment response. Prospective studies should evaluate whether tractography-assisted surgical targeting yields better treatment outcome, and whether one bundle is a superior target compared to the other.

## Data Availability

We are not allowed to share data, since complete anonymization is impossible and the data can be traced back to individual patients.

## ACKNOWLEDGEMENTS

This study was supported by an Innovation Impulse grant (#2015.011) from the Academic Medical Center and a ZonMw grant, nr. 171201008.

